# Thrombocytopenia in critically ill trauma patients is associated with the pattern and duration of post-injury organ dysfunction

**DOI:** 10.1101/2025.03.04.25323139

**Authors:** Andrea Rossetto, Simon Kerner, Ella Ykema, Harriet Allan, Paul Armstrong, Elaine Cole, Paul Vulliamy

**Affiliations:** Centre for Trauma Sciences, Blizard Institute, Queen Mary University of London, London, United Kingdom; Barts Health National Health Service Trust, London, United Kingdom; Centre for Immunobiology, Blizard Institute, Queen Mary University of London, London, United Kingdom

**Keywords:** Critical care, multiple organ failure, thrombocytopenia, trauma

## Abstract

**Background:** Although significant thrombocytopenia is not a common feature of trauma patients in the first hours after injury, little is known about how severe trauma affects platelet count trajectories beyond the initial resuscitation phase, and whether any changes in platelet count are related to clinical outcomes such as the development of post-trauma multiorgan-dysfunction syndrome and mortality.

**Objectives:** To define the incidence, severity and clinical significance of post-injury thrombocytopenia during critical care admission.

**Methods:** Severe trauma patients enrolled in a perpetual cohort study at a single level 1 trauma centre between 2014-2023 and who required critical care admission were included. Thrombocytopenia was classified as mild (100-149×10^9^/L), moderate (50-99×10^9^/L) and severe (<50×10^9^/L). Multivariable regression analyses were used to investigate the drivers of thrombocytopenia and its association with outcomes of organ dysfunction, organ support and mortality.

**Results:** Among the 803 trauma patients investigated, mild, moderate and severe thrombocytopenia occurred in 285 (35%), 290 (36%) and 51 (6%) respectively during their critical care stay. Age, injury severity, shock, admission coagulopathy and total fluid administration within the first 24 hours were all independently associated with the development of moderate-severe thrombocytopenia. Thrombocytopenia of any severity was independently associated with renal and hepatic dysfunction, but not with cardiorespiratory dysfunction or mortality. Severe thrombocytopenia was also independently associated with prolonged need for organ support (OR 2.83, 95%CI 1.07-7.45, p=0.036).

**Conclusions:** Thrombocytopenia is common in injured patients admitted to critical care and severe forms are independently associated with a higher incidence of organ dysfunction and need for organ support.

## INTRODUCTION

Major trauma results in dramatic changes in platelet behaviour, including decreased aggregatory function,^1,2^ loss of response to collagen,^3^ increased surface markers of activation,^3,4^ and procoagulant transformation.^5^ Most previous studies of post-injury platelet biology have focused on how platelets contribute to trauma-induced coagulopathy (TIC) and have largely examined changes in the acute phase after injury.^1,6^ Beyond their roles in haemostasis, platelets are also important contributors to acute inflammatory responses and host defence.^7,8^ Trauma patients who survive the immediate post-injury period are at high risk of multiple organ dysfunction syndromes (MODS), which accounts for much of the critical care morbidity and mortality,^9,10^ but the roles of platelets in the development of post-trauma MODS have not yet been defined in detail.

In addition to early alterations in platelet function after injury, changes in platelet counts during the initial hours after major trauma appear to be clinically important. Although significant thrombocytopenia is not usually a feature of TIC, acute (<24hr) reductions in platelet count even within the normal range are associated with worse outcomes.^6,11^ Beyond 24 hours, less is known about how severe trauma affects platelet count trajectories, and how any changes are related to clinical outcomes. In a recent large multicentre study of mixed intensive care unit (ICU) patients, thrombocytopenia was present in over a third of patients and was associated with increased mortality and resource utilisation.^12^ However, this study only included a small number of injured patients and did not specifically investigate this subgroup. We have recently shown that platelet production dynamics are altered after major trauma, with sustained increases in circulating immature platelets in patients who develop organ failure and thrombosis.^13^ To date, an in-depth study of platelet count trajectories after severe injury, and in particular the significance of thrombocytopenia during post-traumatic critical illness, has not yet been reported.

Our primary objective was to define the incidence, severity and clinical significance of post-injury thrombocytopenia during critical care admission. We hypothesised that thrombocytopenia would be prevalent, and that more severe thrombocytopenia would be associated with higher rates of organ dysfunction, need for organ support, and mortality.

## METHODS

### Study design

Patients were recruited into the perpetual prospective observational cohort study Activation of Coagulation and Inflammation in Trauma (ACIT-II) study (REC reference 07/Q0603/29, ISRCTN12962642). Adult patients requiring activation of the trauma team are screened for inclusion. Exclusion criteria are time from injury >2 hours, administration of >2000ml fluid prehospital, burns >5% body surface area. For the purpose of this study, we included all patients admitted to critical care (high dependency unit or intensive care unit) over a ten-year period between 2014 and 2023, and who had at least one platelet count measurement recorded during the critical care stay. Consent procedures for the ACIT-II study have been described in detail previously.^14^

### Study procedures

Demographic and injury data were recorded prospectively by a trained member of the research team. Participants were followed up daily from the date of injury (designated as day 0) until day 28 post injury, or until death or discharge if this occurred sooner. Variables collected included: daily Sequential Organ Failure Assessment (SOFA) component scores during critical care stay (including platelet count measurements recorded as the haematology component); need for vasopressor support, invasive ventilation and renal replacement therapy (RRT); incidence and type of venous thromboembolic events; and mortality.

Blood samples for research purposes were drawn at admission, at 24 hours and at 72 hours. The immature platelet fraction was measured at these timepoints in a subgroup of the main cohort as part of a separate study using a Sysmex XN-series analyzer according to standard operating procedures, and immature platelet count calculated as previously described.^13^ Rotational thromboelastometry (ROTEM) was performed in citrated whole blood using a Delta instrument according to the manufacturer’s instructions. Base deficit and lactate were measured by point-of-care blood gas analysis.

### Outcomes

Our primary outcome was organ dysfunction and failure, as measured by the SOFA score. In order to exclude the confounding effect of the haematological component (platelet count) we modified the SOFA score as previously described (mSOFA),^15,16^ and considered both the overall mSOFA score and the individual component scores in our analyses. We did not specifically evaluate the central nervous component due to the challenges of accurate daily Glasgow Coma Score measurement in the presence of critical care sedation.^17^ Secondary outcomes were ventilator days, vasopressor days, need for RRT, composite time to complete organ failure recovery (CTCOFR),^18,19^ prolonged organ support, in-hospital mortality; and venous thromboembolism.

### Definitions

Thrombocytopenia was defined as a platelet count of <150×10^9^/L, and further classified as mild (100-149×10^9^/L), moderate (50-99×10^9^/L) and severe (<50×10^9^/L) thrombocytopenia according to previously published definitions.^12^ Patients were subdivided into four groups using these cut-offs according to the lowest count recorded during the critical care stay. Admission injury severity score (ISS) and base deficit were used as markers of injury burden and shock respectively. Coagulopathy was defined as EXTEM A5 <40mm on rotational thromboelastometry.^20^ Massive haemorrhage was defined as the requirement of 10 or more units of red blood cells (RBC) in the first 24 hours. Organ dysfunction and failure in individual organ systems were defined as SOFA component scores of >0 and >2 respectively.^21^ mSOFA was defined as the presence of at least two SOFA components with a score >2, with the exclusion of the haematological component.^15,16^ CTCOFR was defined as the total number of days before mechanical ventilation, vasopressor therapy and RRT were all discontinued, with a score of 0 for those patients who did not require any of these organ support interventions, a score of 15 for those who required any of these interventions for more than 14 days and a score of 16 for those who died at any time during the study period.^18,19^ Prolonged organ disfunction was defined as a CTCOFR of >7 in line with existing definitions.^22^

### Data analysis

Analysis was performed using Prism v10.4.1 (GraphPad Software Inc) and R v4.1.3 (R Core Team). Continuous data are reported as median with interquartile range (IQR) and were compared using the Kruskal-Wallis test with Dunn’s post-test correction for multiple comparisons. Categorical data are reported as number and percentage and compared using Fisher’s exact test or Chi-squared test with Bonferroni correction. No multiple imputation for missing data was performed. A multinomial logistic regression analysis was used to investigate the factors associated with the occurrence of mild, moderate and severe thrombocytopenia. We included the following predictor variables: demographics (sex and age); ISS; shock as measured by base deficit; coagulopathy; and total fluids and blood products administered in the first 24 hours. Multivariable linear and logistic regressions were then used to investigate the association between thrombocytopenia categories and clinical outcomes of organ dysfunction/failure, organ support requirements and mortality, adjusting for the above-mentioned predictor variables and the mechanism of injury. All variables were included in the multivariable regressions independently of their statistical significance in the univariable analysis.^23^ Results are reported as odds ratios (OR) for categorical variables or coefficients for continuous variables with 95% confidence intervals (CI). A two-tailed p-value of <0.05 was considered significant throughout.

## RESULTS

### Patient characteristics

Of the 1800 trauma patients enrolled into the ACIT-II study between 2014 and 2023, the cohort included in this study comprised the 803 patients admitted to critical care and who had at least one measurement of platelet count during the critical care stay. The majority were male (651/803, 81%) and had sustained a blunt mechanism of injury (622/803, 77%) (**table 1**). Patients were severely injured with a median injury severity score of 26 (17-38), a median critical care length of stay of 8 (4-17) days, and an overall mortality of 16% (132/803).

**Table 1.** Characteristics of the study cohort stratified by the severity of thrombocytopenia in critical care.

|  | Overall<br>(n = 803) | No<br>Thrombocytopenia<br>(n = 177) | Mild<br>Thrombocytopenia<br>(n = 285) | Moderate<br>Thrombocytopenia<br>(n = 290) | Severe<br>Thrombocytopenia<br>(n = 51) |
| --- | --- | --- | --- | --- | --- |
| **Admission characteristics** |  |  |  |  |  |
| Sex, male | 651 (81.1) | 145 (81.9) | 229 (80.4) | 240 (82.8) | 37 (72.5) |
| Age, years | 37 (25-55) | 37 (26-53) | 38 (26-58) | 35 (24-53) | 36 (26-53) |
| Glasgow coma score | 11 (5-15) | 11 (6-15) | 11 (5-15) | 11 (6-15) | 10 (3-14) |
| Base deficit, mEq/L | 4 (1-9) | 2 (0-5) | 3 (1-7)* | 6 (3-11)*§ | 10 (5-18)*§† |
| SBP, mmHg | 117 (92-139) | 127 (106-146) | 119 (95-140) | 110 (89-134)*§ | 97 (75-126)*§ |
| INR >1.2 | 154 (23.7) | 12 (8.3) | 47 (20.1)* | 81 (34.3)*§ | 14 (38.9)* |
| EXTEM A5, mm | 40 (34-45) | 45 (40-49) | 41 (36-46)* | 36 (31-42)*§ | 32 (22-41)*§ |
| **Injury characteristics** |  |  |  |  |  |
| Mechanism of injury | 622 (77.5) | 138 (78.0) | 220 (77.2) | 223 (76.9) | 41 (80.4) |
| Injury severity score | 26 (17-38) | 24 (16-30) | 26 (17-35)* | 29 (19-41)* | 32 (25-43)*§ |
| AIS Head/Neck ≥ 3 | 396 (50.2) | 94 (54.7) | 153 (54.3) | 126 (44.1) | 23 (46.9) |
| AIS Face ≥ 3 | 32 (4.1) | 6 (3.5) | 10 (3.6) | 13 (4.5) | 3 (6.1) |
| AIS Thorax ≥ 3 | 423 (53.2) | 85 (48.6) | 152 (53.7) | 165 (57.5) | 21 (42.0) |
| AIS Abdomen ≥ 3 | 185 (23.5) | 27 (15.5) | 55 (19.7) | 87 (30.6)*§ | 16 (32.0) |
| AIS Extremity ≥ 3 | 280 (35.4) | 27 (15.7) | 79 (28.0)* | 145 (50.3)*§ | 29 (58.0)*§ |
| AIS External ≥ 3 | 19 (2.5) | 1 (0.6) | 10 (3.7) | 5 (1.8) | 3 (6.7) |
| **Fluids and BP in first 24hr** |  |  |  |  |  |
| Crystalloids, L | 3.00 (2.00-4.20) | 2.03 (1.32-3.38) | 3.00 (2.00-4.01)* | 3.18 (2.20-4.5)*§ | 3.40 (2.50-5.00)* |
| Red blood cells, units | 2 (0-6) | 0 (0-2) | 2 (0-4)* | 4 (2-8)*§ | 9 (5-17)*§† |
| Massive haemorrhage | 87 (11.1) | 3 (1.7) | 9 (3.2) | 52 (18.1)*§ | 23 (46.9)*§† |
| Fresh frozen place, units | 1 (0-5) | 0 (0-0) | 0 (0-4)* | 4 (0-8)*§ | 8 (4-14)*§† |
| Platelets, pools | 0 (0-1) | 0 (0-0) | 0 (0-1)* | 1 (0-2)*§ | 1 (1-3)*§† |
| Cryoprecipitate, pools | 0 (0-2) | 0 (0-0) | 0 (0-1)* | 2 (0-3)*§ | 3 (1-6)*§† |

Data presented as median (interquartile range) or count (percentage).
\* p <0.05 when compared to No Thrombocytopenia; § p <0.05 when compared to Mild Thrombocytopenia; † p <0.05 when compared to Moderate Thrombocytopenia.
SBP, systolic blood pressure; INR; international normalised ratio; AIS, abbreviated injury severity score; BP, blood products.

### Platelet count trajectories after injury

On arrival in the emergency department, thrombocytopenia was present in 119/803 patients (15%), most of whom had mild reductions in platelet count only. During the critical care stay, the incidence of thrombocytopenia rose to 78% (626/803), with markedly higher rates of moderate (36.1% vs 3.1%, p<0.001) and severe (6.4% vs 0.6%, p<0.001) thrombocytopenia compared to admission (**figure 1A**). Overall, mild, moderate and severe thrombocytopenia occurred in 285 (35%), 290 (36%) and 51 (6%) patients respectively during the critical care stay. Longitudinal platelet counts trajectories were similar in these subgroups with an initial decline in platelet count, reaching a nadir between 48-72 hours after injury irrespective of the presence or severity of thrombocytopenia (**figure 1B**). In patients with mild and moderate thrombocytopenia, platelet counts recovered by day 7 before rising to supra-normal levels by day 14 and beginning to return to normal thereafter. Patients with severe thrombocytopenia showed a similar trajectory, but with a slower recovery, lower peak, and persistently lower count throughout the critical care stay.

**Figure 1:**
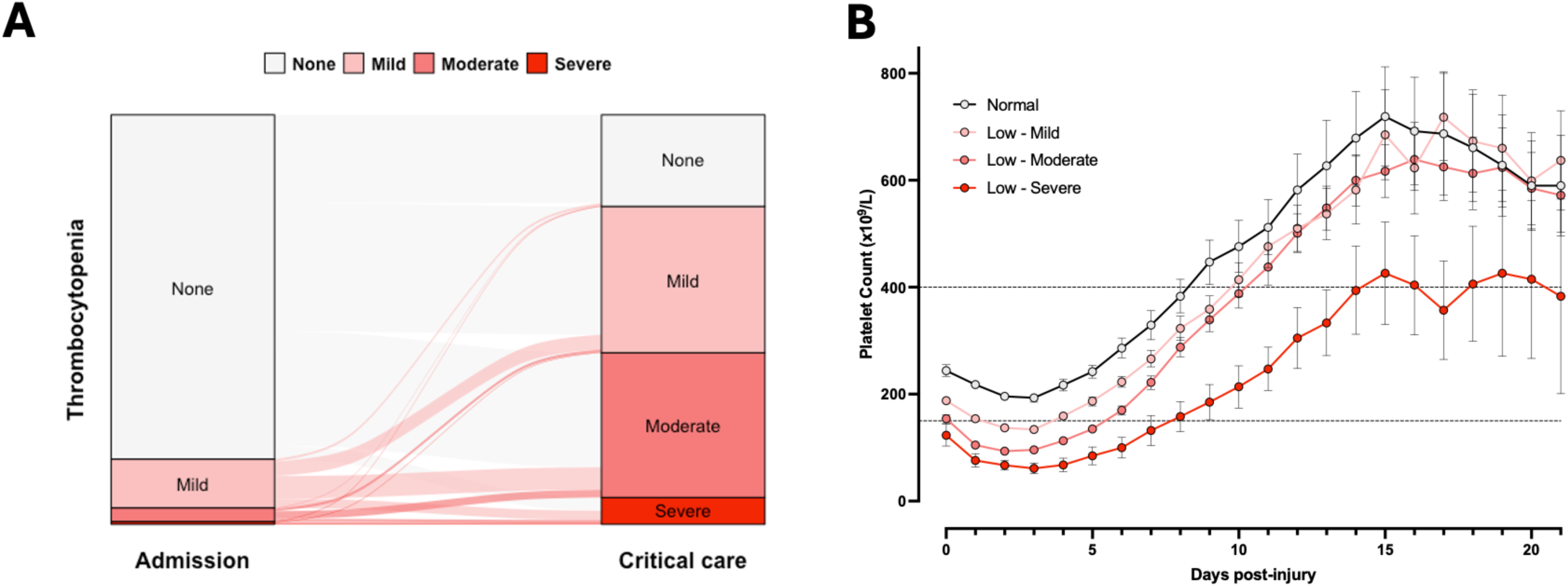
Platelet count trajectories after injury. **A:** Sankey plot depicting thrombocytopenia patterns on arrival in the emergency department (left column) and during critical care stay (right column). None, no thrombocytopenia; mild (100-149×10^9^/L), moderate (50-99×10^9^/L) and severe (<50×10^9^/L). **B:** Platelet counts over time in patients grouped by lowest platelet count during critical care stay. Mean with 95% confidence intervals. Dashed lines denote normal range.

### Factors associated with post-injury thrombocytopenia

We next examined the clinical characteristics of patients stratified by severity of thrombocytopenia during their critical care admission. Patients with moderate and severe thrombocytopenia were more severely injured, shocked and coagulopathic on arrival compared to those with mild and no thrombocytopenia (**table 1**). However, there were no differences in mechanism of injury, patient age, or incidence of traumatic brain injury across the four groups. Volumes of blood products (including platelet transfusions) and crystalloid administered in the first 24 hours after injury were higher with increasing degree of thrombocytopenia. However, the haematocrit and haemoglobin concentrations over the first 72 hours were similar irrespective of the degree of thrombocytopenia (**supplemental figure 1**). On multivariable analysis, we found that older age, injury severity, degree of shock, admission coagulopathy and total fluid administration within the first 24 hours were all independently associated with development of both moderate and severe thrombocytopenia (**table 2**).

**Table 2.** Multinomial logistic regression analysis for severity of thrombocytopenia in critical care.

|  | Severity | OR 95% CI | p | Adj. OR 95% CI | Adj. p |
| --- | --- | --- | --- | --- | --- |
| Sex, male | Mild | 0.80 (0.47 to 1.36) | 0.401 | 0.95 (0.54 to 1.66) | 0.853 |
|  | Moderate | 0.90 (0.53 to 1.54) | 0.700 | 1.04 (0.56 to 1.92) | 0.913 |
|  | Severe | 0.40 (0.18 to 0.87) | 0.020 | 0.44 (0.18 to 1.08) | 0.073 |
| Age, years | Mild | 1.01 (1.00 to 1.02) | 0.119 | 1.02 (1.00 to 1.03) | 0.007 |
|  | Moderate | 1.00 (0.99 to 1.01) | 0.724 | 1.02 (1.00 to 1.03) | 0.016 |
|  | Severe | 1.00 (0.98 to 1.02) | 0.968 | 1.02 (1.00 to 1.05) | 0.040 |
| Injury severity score | Mild | 1.02 (1.00 to 1.04) | 0.015 | 1.01 (1.00 to 1.03) | 0.111 |
|  | Moderate | 1.04 (1.03 to 1.06) | <0.001 | 1.02 (1.01 to 1.04) | 0.012 |
|  | Severe | 1.05 (1.03 to 1.08) | <0.001 | 1.03 (1.00 to 1.06) | 0.033 |
| Base deficit, mEq/L | Mild | 1.06 (1.01 to 1.11) | 0.015 | 1.02 (0.97 to 1.07) | 0.479 |
|  | Moderate | 1.15 (1.10 to 1.20) | <0.001 | 1.05 (1.00 to 1.10) | 0.053 |
|  | Severe | 1.24 (1.17 to 1.30) | <0.001 | 1.11 (1.04 to 1.18) | 0.001 |
| EXTEM <40 mm | Mild | 2.10 (1.32 to 3.33) | 0.002 | 1.84 (1.14 to 2.98) | 0.013 |
|  | Moderate | 6.99 (4.40 to 11.1) | <0.001 | 4.60 (2.77 to 7.65) | <0.001 |
|  | Severe | 7.61 (3.57 to 16.2) | <0.001 | 2.99 (1.23 to 7.29) | 0.016 |
| Total fluids and BP in first 24hr, L | Mild | 1.22 (1.12 to 1.33) | <0.001 | 1.21 (1.10 to 1.33) | <0.001 |
|  | Moderate | 1.51 (1.38 to 1.64) | <0.001 | 1.42 (1.29 to 1.56) | <0.001 |
|  | Severe | 1.70 (1.53 to 1.87) | <0.001 | 1.57 (1.40 to 1.75) | <0.001 |

R<sup>2</sup> = 0.08/0.32/0.51. Cases per variable = 25.0/25.0/7.0.
OR, odds ratio; CI, confidence interval; Adj., adjusted. BP, blood products.

We also examined immature platelet metrics in a subgroup of 92 patients (no thrombocytopenia, n=19; mild, n=29; moderate, n=39; severe, n=5); characteristics of this subgroup are reported in **supplemental table 1**. We found no statistically significant differences in either IPF or IPC at admission, 24 hours or 72 hours in patients stratified according to thrombocytopenia severity (**supplemental figure 2)**.

### Association between thrombocytopenia severity and clinical outcomes

Patients with moderate and severe thrombocytopenia had significantly higher mSOFA (**figure 2A**) and CTCOFR scores (**figure 2B**) compared to those without thrombocytopenia. SOFA component scores for cardiovascular, renal and hepatic dysfunction were also consistently higher throughout critical care stay in patients with severe thrombocytopenia, while respiratory scores were similar across the four groups (**supplemental figure 3**). Organ support requirements were significantly higher in patients with moderate and severe thrombocytopenia, with higher ventilator days (**figure 2C**), vasopressor days (**figure 2D**) and proportions requiring RRT (**figure 2E**). Rates of VTE rose in a linear manner with increasing severity of thrombocytopenia although this trend did not reach statistical significance (**figure 2F**). Overall mortality was significantly higher in patients with moderate and severe thrombocytopenia compared to those with no thrombocytopenia (**figure 2G-H**).

**Figure 2:**
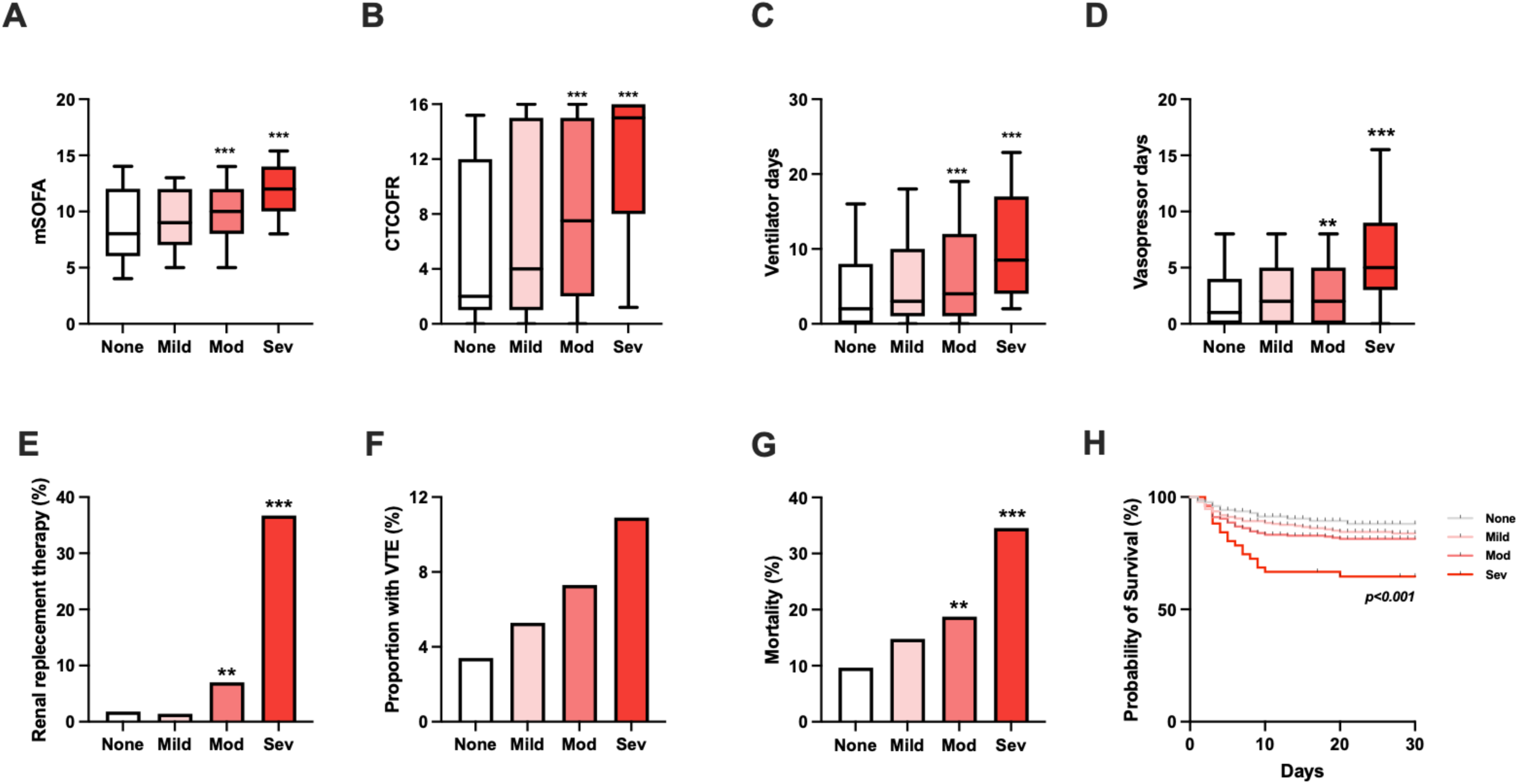
Association between severity of thrombocytopenia with clinical outcomes. **A**: Modified sequential organ failure assessment (SOFA) score, calculated as sum of all individual component scores except platelet count. **B**: Composite time to complete organ failure (CTCOFR). **C**: Ventilator days. **D**: Days requiring vasopressor support. **E**: Renal replacement therapy use. **F**: Venous thromboembolism. **G**: In-hospital mortality. **H**: Kaplan-Meier curves for survival. P-value derived using log-rank test. **p<0.01, ***p<0.001 vs non-thrombocytopenia, Kruskal-Wallis test with Dunn’s post-test correction. Box-Whisker plots depict 10th-90^th^ percentiles.

To further investigate the relationship between thrombocytopenia severity and outcome, we constructed a series of regression models. On univariable analysis, both moderate and severe thrombocytopenia were associated with respiratory, cardiovascular, renal and hepatic dysfunction, as well as higher odds of prolonged organ support and mortality (**figure 3A**). On multivariable analysis, mild, moderate and severe thrombocytopenia remained independently associated with renal (mild: OR 1.96, 95%CI 1.17-3.29, p=0.011; moderate: OR 2.82, 95%CI 1.61-4.93, p<0.001; severe: OR 8.33, 95%CI 2.93-23.6, p=<0.001), and hepatic dysfunction (mild: OR 2.13, 95%CI 1.34-3.39, p=0.001; moderate: OR 3.38, 95%CI 2.01-5.69, p<0.001; severe: OR 6.34, 95%CI 2.26-17.8, p=<0.001), but not with cardiorespiratory dysfunction or mortality after adjustment for relevant confounders (**figure 3B and supplemental table 2**). Severe thrombocytopenia was also independently associated with prolonged need for organ support (OR 2.83, 95%CI 1.07-7.45, p=0.036), CTCOFR (coefficient 2.27, 95%CI 0.11-4.44, p=0.040), ventilator days (coefficient 0.85, 95%CI 0.14-1.55, p=0.019), vasopressor days (coefficient 0.70, 95%CI 0.21-1.19, p=0.005) and need for RRT (OR 9.22, 95%CI 1.42-59.8, p=0.020) on multivariable analysis (**supplemental table 3**).

**Figure 3:**
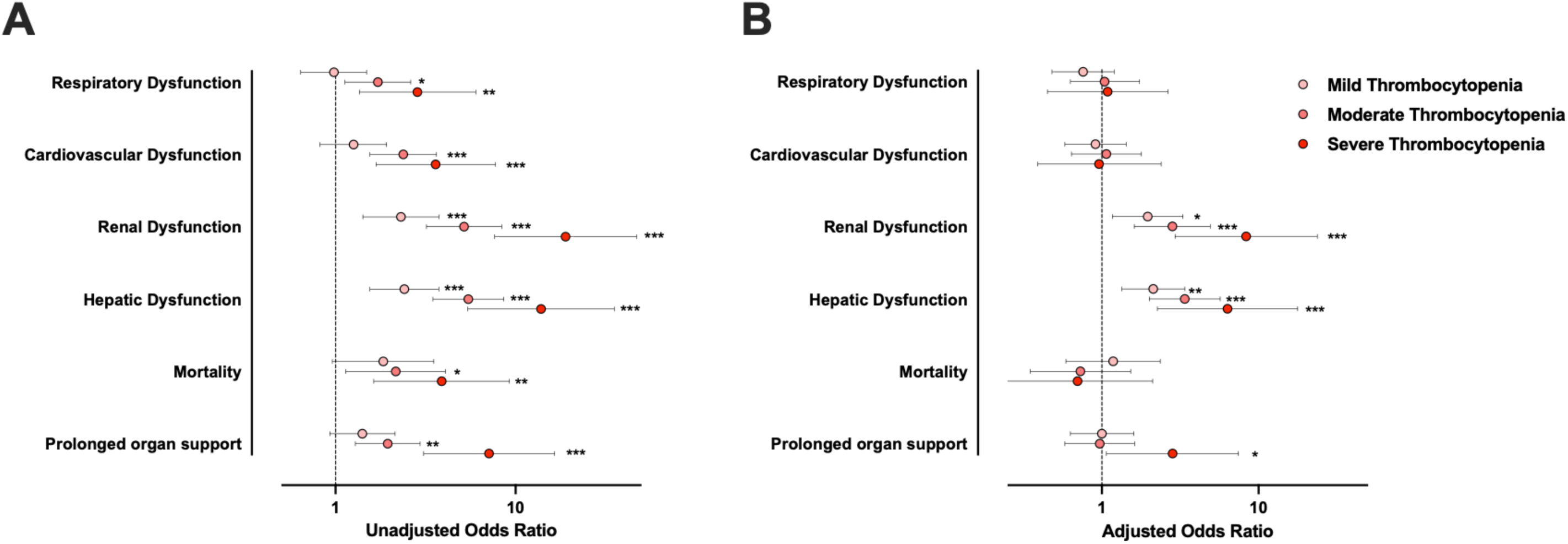
Multivariable regression analysis for organ dysfunction, mortality and prolonged organ support. **A:** Univariable models. **B:** Multivariable models after adjusting for age, sex, injury severity score, mechanism of injury, admission base deficit, presence of coagulopathy, total fluid volumes administered within 24 hours. Full details of all models are reported in supplemental table 2. Organ dysfunction defined as a SOFA component score >0 for the cardiovascular, renal and hepatic components and >2 for the respiratory component. Prolonged organ support defined as a composite time to complete organ failure recovery (CTCOFR) score >7. Circles are odds ratios relative to no thrombocytopenia. Error bars represent 95% confidence intervals. *p<0.05, **p<0.01, ***p<0.001.

## DISCUSSION

In this study of over 800 critically ill major trauma patients, we report the clinical relevance and drivers of post-injury thrombocytopenia. Our central findings are that some degree of thrombocytopenia is extremely common in critical care trauma patients; that moderate-severe depletion of platelets is associated with both endogenous and iatrogenic factors; and third, that severe thrombocytopenia is independently associated with a higher incidence of organ dysfunction and need for organ support, but not with mortality. These findings suggest a process of increased platelet destruction, sequestration, dilution and/or consumption after injury which is associated with progression to organ failure. The mechanisms involved in this process are not clear and warrant further mechanistic investigation.

More than three-quarters of injured patients in this study developed some degree of thrombocytopenia, a significantly higher proportion than in previous studies of critically ill patients. In the recent PLOT-ICU study,^12^ the overall incidence of thrombocytopenia in a mixed ICU population (primarily with cardiorespiratory failure and/or sepsis) was 43%, and in a large study of neonatal patients post-cardiac surgery the incidence was 34%.^24^ The dynamics of thrombocytopenia in these populations were also different to the post-injury response, with post-cardiac surgery patients recovering steadily from a nadir on post-operative day 1, and the mixed critically ill population in PLOT-ICU having higher rates of admission thrombocytopenia compared to trauma patients. Collectively, this suggests that although platelet depletion appears to be a common feature in multiple acutely ill patient populations, the incidence and potential underlying mechanisms vary across disease states.

We found that post-injury thrombocytopenia occurs due to a combination of age, injury-related factors and the dilutional effect of fluid resuscitation given in the immediate post-injury period. Both injury load and degree of shock were independently associated with the development of moderate-severe thrombocytopenia. Platelet depletion in critical illness is often variously attributed to consumption, sequestration and/or enhanced clearance, but direct evidence for these processes in trauma patients is lacking. However, given our observation here that early immature platelet metrics do not correlate with thrombocytopenia, it appears that peripheral depletion of platelets rather than bone marrow failure is primarily responsible. Given that both tissue injury and systemic hypoperfusion lead to release of damage-associated molecular patterns (DAMPs),^25^ many of which are known to induce thrombocytopenia and modulate platelet activity,^5,26^ we postulate that DAMP-mediated platelet depletion could represent the central mechanism, although clearly further study in this area is required to corroborate this.

In contrast to the aforementioned studies of thrombocytopenia in other patient populations,^12,24^ we did not find an independent association between thrombocytopenia and mortality. However, we found that severely low platelet counts were associated with prolonged need for organ support, and that thrombocytopenia overall was associated with liver and renal (although not cardio-respiratory) dysfunction. Severe thrombocytopenia in septic patients has been shown to result in specific perturbations in immune function, with increased cytokine levels, endothelial cell activation and impaired vascular integrity.^15^ Consistent with this, our findings suggest that marked depletion of platelets after injury confers a greater risk of more severe and more prolonged organ failure, with implications for resource utilisation and patient morbidity. The immunological mechanisms that may be involved in the trauma setting remain to be elucidated.

This study is limited by its single centre observational design, which limits generalisability and potential for causal inference. In addition, data were lacking on fluid volumes administered beyond 24 hours, incidence of nosocomial infection, and immature platelet metrics for the full cohort. We did not specifically evaluate the effect of platelet transfusions given after resuscitation on platelet counts, and this remains an important area for further study. Finally, because we use the SOFA score to quantify organ dysfunction, we were not able to directly examine the relationship between platelet counts and MODS risk because the complete SOFA score calculation includes platelet count. To overcome this limitation we focused on organ support, which represents a more direct measure of the resource utilisation associated with organ dysfunction.

In summary, this study provides a detailed insight into the factors associated with post-injury thrombocytopenia and its clinical relevance. In contrast with other patient populations, the severity of thrombocytopenia on critical care is not independently associated with mortality but is linked to the pattern of organ dysfunction and the overall requirement for organ support. The main factors associated with development of thrombocytopenia are age, injury burden, degree of hypoperfusion at admission, and fluid resuscitation volumes. Further work is required to interrogate the underlying mechanisms responsible for post-injury thrombocytopenia and the relationship between platelet depletion and altered immunological responses.

## AUTHOR CONTRIBUTIONS

A.R., S.K., E.C. and P.V. designed the study. A.R., S.K., E.Y. and P.V. managed and analysed the data. All authors contributed to data interpretation. A.R., S.K., E.C. and P.V. wrote the manuscript. All authors critically revised the intellectual content and approved the final version of the manuscript for publication.

## DATA AVAILABILITY STATEMENT

The data that support the findings of this study are available from the corresponding author, P.V., upon reasonable request.

## Funding information

H.A. received funding from the British Heart Foundation (FS/IPBSRF/23/27090). P.A. received funding from the British Heart Foundation (RG/19/8/34500). P.V. received funding from Wellcome (Early Career Award, 309168/Z/24/Z). The Centre for Trauma Science (Blizard Institute, Queen Mary University of London, London, United Kingdom) receives departmental support for rotational thromboelastometry reagents and equipment from TEM International GmbH (Munich, Germany).

## Conflict of interest

The authors declare that they have no conflicts of interest.

## Supporting information

Supplemental File and Strobe Checklist

## Data Availability

All data produced in the present study are available upon reasonable request to the authors.

## Notes

### Competing Interest Statement

The authors have declared no competing interest.

### Author Declarations

The East London and The City Research Ethics Committee gave ethical approval for this work (07/Q0603/29)

