## Supplemental File and Strobe Checklist for "Thrombocytopenia in critically ill trauma patients is associated with the pattern and duration of post-injury organ dysfunction"

**Supplemental file content:**

- **Supplemental Table 1.** Characteristics of the immature platelet fraction (IPF) cohort stratified by severity of thrombocytopenia in critical care.
- **Supplemental Table 2.** Multivariable logistic regression analysis for mortality, organ dysfunction/failure and organ failure resolution.
- **Supplemental Table 3.** Multivariable linear regression analysis for organ failure resolution and organ support.
- **Supplemental Figure 1.** Haematocrit and haemoglobin levels over time stratified by severity of thrombocytopenia.
- **Supplemental Figure 2.** I immature platelet fraction (IPF) and immature platelet count (IPC) levels over time stratified by severity of thrombocytopenia.
- **Supplemental Figure 3.** Organ injury score trajectories stratified by severity of thrombocytopenia.

**Supplemental Table 1. Characteristics of the IPF cohort stratified by the severity of thrombocytopenia in critical care**

| | Overall | No Thrombocytopenia | Mild Thrombocytopenia | Moderate Thrombocytopenia | Severe Thrombocytopenia |
|---|---|---|---|---|---|
| | (n = 92) | (n = 19) | (n = 29) | (n = 39) | (n = 5) |
| **Admission characteristics** | | | | | |
| Sex, male | 75 (81.5) | 15 (78.9) | 22 (75.9) | 33 (84.6) | 5 (100.0) |
| Age, years | 35 (22-53) | 33 (23-44) | 32 (21-56) | 40 (23-53) | 35 (25-41) |
| Glasgow coma score | 12 (4-15) | 14 (7-15) | 14 (8-15) | 8 (3-14) | 3 (3-3) |
| Base deficit, mEq/L | 5 (2-9) | 3 (0-7) | 2 (1-7) | 7 (4-15) | 5 (2-17) |
| SBP, mmHg | 114 (98-131) | 117 (96-139) | 118 (107-127) | 111 (101-131) | 93 (59-127) |
| INR >1.2 | 21 (29.2) | 3 (17.6) | 6 (25.0) | 11 (36.7) | 1 (100.0) |
| EXTEM A5, mm | 37 (30-42) | 39 (36-45) | 39 (32-44) | 35 (29-39) | 19 (12-31) |
| **Injury characteristics** | | | | | |
| Mechanism of injury | 65 (70.7) | 13 (68.4) | 17 (58.6) | 32 (82.1) | 3 (60.0) |
| Injury severity score | 29 (21-38) | 21 (13-27) | 25 (17-33) | 36 (25-45) | 30 (30-35) |
| AIS Head/Neck ≥ 3 | 40 (45.5) | 7 (46.7) | 11 (37.9) | 19 (48.7) | 3 (60.0) |
| AIS Face ≥ 3 | 4 (4.5) | 1 (6.3) | 1 (3.4) | 2 (5.1) | 0 (0.0) |
| AIS Thorax ≥ 3 | 51 (55.4) | 9 (47.4) | 14 (48.3) | 27 (69.2) | 1 (20.0) |
| AIS Abdomen ≥ 3 | 33 (37.1) | 4 (23.5) | 13 (44.8) | 15 (39.5) | 1 (20.0) |
| AIS Extremity ≥ 3 | 30 (34.1) | 4 (25.0) | 5 (17.2) | 19 (50.0) | 2 (40.0) |
| AIS External ≥ 3 | 1 (1.1) | 0 (0.0) | 0 (0.0) | 0 (0.0) | 1 (20.0) |
| **Fluids and BP in first 24hr** | | | | | |
| Crystalloids, L | 2.23 (1.32-3.83) | 1.53 (1.32-3.00) | 2.00 (1.10-3.30) | 2.53 (1.35-4.25) | 4.00 (3.00-6.45) |
| Red blood cells, units | 3 (1-7) | 2 (0-4) | 3 (1-4) | 5 (2-9) | 7 (0-33) |
| Massive haemorrhage | 9 (10.8) | 0 (0.0) | 1 (4.0) | 7 (18.4) | 1 (33.3) |
| Fresh frozen place, units | 2 (0-6) | 1 (0-3) | 1 (0-4) | 4 (0-8) | 11 (0-27) |
| Platelets, pools | 0 (0-1) | 0 (0-1) | 0 (0-1) | 1 (0-2) | 1 (0-3) |
| Cryoprecipitate, pools | 0 (0-2) | 0 (0-2) | 1 (0-2) | 2 (0-4) | 0 (0-11) |
| **Outcomes** | | | | | |
| mSOFA | 56 (62.2) | 8 (47.1) | 13 (44.8) | 31 (79.5) | 4 (80.0) |
| CTCOFR, days | 9 (1-16) | 3 (1-16) | 3 (1-9) | 14 (5-16) | 16 (16-16) |
| Ventilator days | 3 (1-10) | 2 (0-11) | 2 (1-8) | 6 (2-13) | 7 (3-9) |
| Vasopressor days | 2 (0-5) | 1 (0-2) | 1.0 (0-3) | 3 (2-6) | 6 (3-7) |
| RRT | 7 (7.7) | 1 (5.6) | 0 (0.0) | 4 (10.3) | 2 (40.0) |
| Venous thromboembolism | 7 (7.6) | 0 (0.0) | 1 (3.4) | 5 (12.8) | 1 (20.0) |
| Mortality | 26 (28.3) | 5 (26.3) | 3 (10.3) | 14 (35.9) | 4 (80.0) |

Data presented as median (interquartile range) or count (percentage).
IPF, immature platelet fraction; SBP, systolic blood pressure; INR; international normalised ratio; AIS, abbreviated injury severity score; BP, blood products; mSOFA, modified sequential organ failure assessment score; CTCOFR, Composite time to complete organ failure; RRT, renal replacement therapy.

**Supplemental Table 2. Multivariable regression analysis for mortality, organ dysfunction/failure and organ failure resolution.**

| | Respiratory failure | | Cardiovascular dysfunction/failure | | Renal dysfunction/failure | | Hepatic dysfunction/failure | | Mortality | | CTCOFR | |
|---|---|---|---|---|---|---|---|---|---|---|---|---|
| | Adj. OR (95% CI) | p | Adj. OR (95% CI) | p | Adj. OR (95% CI) | p | Adj. OR (95% CI) | p | Adj. OR (95% CI) | p | Adj. OR (95% CI) | p |
| Sex, males | 1.57 (1.03 to 2.40) | **0.035** | 0.61 (0.29 to 1.32) | 0.210 | 3.57 (2.19 to 5.83) | **<0.001** | 0.95 (0.62 to 1.45) | 0.800 | 0.86 (0.51 to 1.47) | 0.592 | 1.08 (0.70 to 1.67) | 0.732 |
| Age, years | 1.02 (1.01 to 1.03) | **0.002** | 1.01 (0.99 to 1.02) | 0.466 | 1.01 (1.00 to 1.02) | 0.130 | 0.99 (0.98 to 1.00) | **0.048** | 1.03 (1.02 to 1.05) | **<0.001** | 1.02 (1.01 to 1.03) | **<0.001** |
| Mechanism of injury, blunt | 3.24 (1.97 to 5.31) | **<0.001** | 2.34 (1.21 to 4.50) | **0.011** | 0.92 (0.56 to 1.52) | 0.750 | 1.10 (0.68 to 1.78) | 0.699 | 1.64 (0.77 to 3.48) | 0.199 | 2.77 (1.64 to 4.67) | **<0.001** |
| Injury severity score | 1.03 (1.01 to 1.04) | **<0.001** | 1.04 (1.01 to 1.06) | **0.004** | 1.02 (1.00 to 1.03) | **0.019** | 1.02 (1.00 to 1.03) | **0.020** | 1.03 (1.01 to 1.05) | **<0.001** | 1.06 (1.04 to 1.07) | **<0.001** |
| Base deficit, mEq/L | 1.05 (1.02 to 1.08) | **0.003** | 1.01 (0.96 to 1.05) | 0.781 | 1.08 (1.05 to 1.12) | **<0.001** | 0.99 (0.96 to 1.02) | 0.652 | 1.05 (1.01 to 1.09) | **0.006** | 1.05 (1.02 to 1.08) | **0.004** |
| EXTEM <40 mm | 0.99 (0.69 to 1.43) | 0.969 | 1.33 (0.75 to 2.35) | 0.324 | 1.07 (0.74 to 1.57) | 0.712 | 0.99 (0.69 to 1.44) | 0.970 | 2.49 (1.50 to 4.15) | **<0.001** | 1.61 (1.10 to 2.34) | **0.013** |
| Total Fluids and BP in 24h, L | 1.06 (1.00 to 1.11) | **0.035** | 1.17 (1.05 to 1.30) | **0.005** | 1.07 (1.02 to 1.14) | **0.011** | 1.13 (1.07 to 1.20) | **<0.001** | 1.05 (1.00 to 1.11) | **0.044** | 1.02 (0.97 to 1.07) | 0.387 |
| Thrombocytopenia | | | | | | | | | | | | |
| Mild | 0.76 (0.48 to 1.20) | 0.235 | 1.07 (0.57 to 1.98) | 0.835 | 1.96 (1.17 to 3.29) | **0.011** | 2.13 (1.34 to 3.39) | **0.001** | 1.18 (0.59 to 2.36) | 0.633 | 1.00 (0.63 to 1.60) | 0.998 |
| Moderate | 1.04 (0.63 to 1.74) | 0.871 | 1.21 (0.56 to 2.63) | 0.621 | 2.82 (1.61 to 4.93) | **<0.001** | 3.38 (2.01 to 5.69) | **<0.001** | 0.73 (0.35 to 1.53) | 0.403 | 0.97 (0.58 to 1.62) | 0.893 |
| Severe | 1.09 (0.45 to 2.64) | 0.843 | 2.15 (0.26 to 18.1) | 0.481 | 8.33 (2.93 to 23.6) | **<0.001** | 6.34 (2.26 to 17.8) | **<0.001** | 0.70 (0.23 to 2.11) | 0.529 | 2.83 (1.07 to 7.45) | **0.036** |

Mortality: $R^2$ = 0.15, cases per variable = 13.8. Respiratory failure: $R^2$ = 0.11, cases per variable = 38.8, survivors only. Cardiovascular dysfunction/failure: $R^2$ = 0.12, cases per variable = 9.3, survivors only. Renal dysfunction/failure: $R^2$ = 0.18, cases per variable = 36.9, survivors only. Hepatic dysfunction/failure: $R^2$ = 0.13, cases per variable = 36.3, survivors only. CTCOFR: $R^2$ = 0.18, cases per variable = 41.9. CTCOFR, composite time to complete organ failure resolution; Adj., adjusted. OR, odds ratio; CI, confidence interval; BP, blood product.

**Supplemental Table 3. Multivariable regression analysis for organ failure resolution and organ support.**

| | CTCOFR | | | Ventilator days | | | Vasopressor days | | | RRT | | |
|---|---|---|---|---|---|---|---|---|---|---|---|---|
| | Adj. Coef. | (95% CI) | p | Adj. Coeff. | (95% CI) | p | Adj. Coeff. | (95% CI) | p | OR | (95% CI) | p |
| Sex, males | -0.17 | (-1.24 to 0.89) | 0.750 | -0.08 | (-0.41 to 0.25) | 0.616 | 0.00 | (-0.24 to 0.24) | 0.972 | 3.37 | (0.76 to 15.0) | 0.110 |
| Age, years | 0.06 | (0.03 to 0.08) | **<0.001** | 0.01 | (0.00 to 0.02) | **0.033** | 0.01 | (0.00 to 0.01) | **0.019** | 0.99 | (0.96 to 1.02) | 0.358 |
| Mechanism of injury, blunt | 2.26 | (1.09 to 3.44) | **<0.001** | 0.37 | (0.02 to 0.72) | **0.038** | 0.36 | (0.10 to 0.61) | **0.006** | 2.66 | (0.61 to 11.6) | 0.194 |
| Injury severity score | 0.15 | (0.12 to 0.19) | **<0.001** | 0.05 | (0.04 to 0.06) | **<0.001** | 0.03 | (0.02 to 0.04) | **<0.001** | 1.05 | (1.01 to 1.08) | **0.005** |
| Base deficit, mEq/L | 0.12 | (0.05 to 0.19) | **0.001** | 0.02 | (-0.00 to 0.04) | 0.094 | 0.00 | (-0.02 to 0.02) | 0.868 | 1.03 | (0.96 to 1.10) | 0.409 |
| EXTEM <40 mm | 1.44 | (0.52 to 2.36) | **0.002** | 0.19 | (-0.09 to 0.47) | 0.175 | 0.15 | (-0.05 to 0.35) | 0.149 | 0.74 | (0.26 to 2.13) | 0.575 |
| Total Fluids and BP in 24h, L | 0.07 | (-0.04 to 0.19) | 0.194 | 0.03 | (-0.01 to 0.07) | 0.175 | 0.03 | (-0.00 to 0.06) | 0.071 | 1.16 | (1.05 to 1.28) | **0.004** |
| Thrombocytopenia | | | | | | | | | | | | |
| Mild | 0.26 | (-0.89 to 1.41) | 0.655 | 0.04 | (-0.30 to 0.38) | 0.810 | -0.04 | (-0.29 to 0.20) | 0.726 | 0.76 | (0.12 to 4.84) | 0.775 |
| Moderate | 0.30 | (-0.96 to 1.55) | 0.643 | 0.09 | (-0.29 to 0.47) | 0.634 | 0.04 | (-0.23 to 0.32) | 0.757 | 1.68 | (0.31 to 9.00) | 0.547 |
| Severe | 2.27 | (0.11 to 4.44) | **0.040** | 0.85 | (0.14 to 1.55) | **0.019** | 0.70 | (0.21 to 1.19) | **0.005** | 9.22 | (1.42 to 59.8) | **0.020** |

CTCOFR: $R^2$ = 0.27, cases per variable = 85.5. Ventilator days: $R^2$ = 0.19, cases per variable = 71, survivors only. Vasopressors days: $R^2$ = 0.21, cases per variable = 69.5, survivors only. RRT days: $R^2$ = 0.15, cases per variable = 3.3, survivors only. CTCOFR, composite time to complete organ failure resolution; RRT, renal replacement therapy; Adj., adjusted; Coef., coefficient; CI, confidence interval; OR, odds ratio; BP, blood product.

**Supplemental Figure 1.** Haematocrit (A) and haemoglobin levels (B) over time stratified by the severity of thrombocytopenia. Bars indicate median with standard deviation.

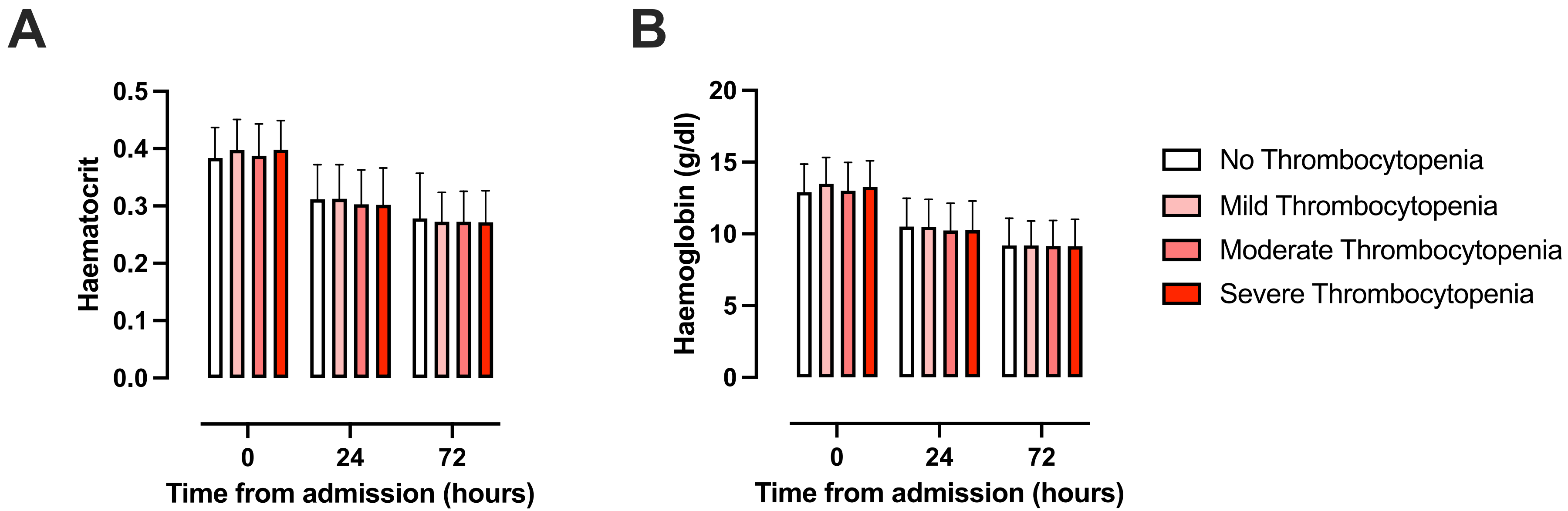

**Supplemental Figure 2.** Immature platelet fraction (IPF) and immature platelet count (IPC) levels over time stratified by the severity of thrombocytopenia. None, no thrombocytopenia; Mild, mild thrombocytopenia; Mod, moderate thrombocytopenia; Sev, Severe thrombocytopenia. Whiskers represent 10th-90th percentiles.

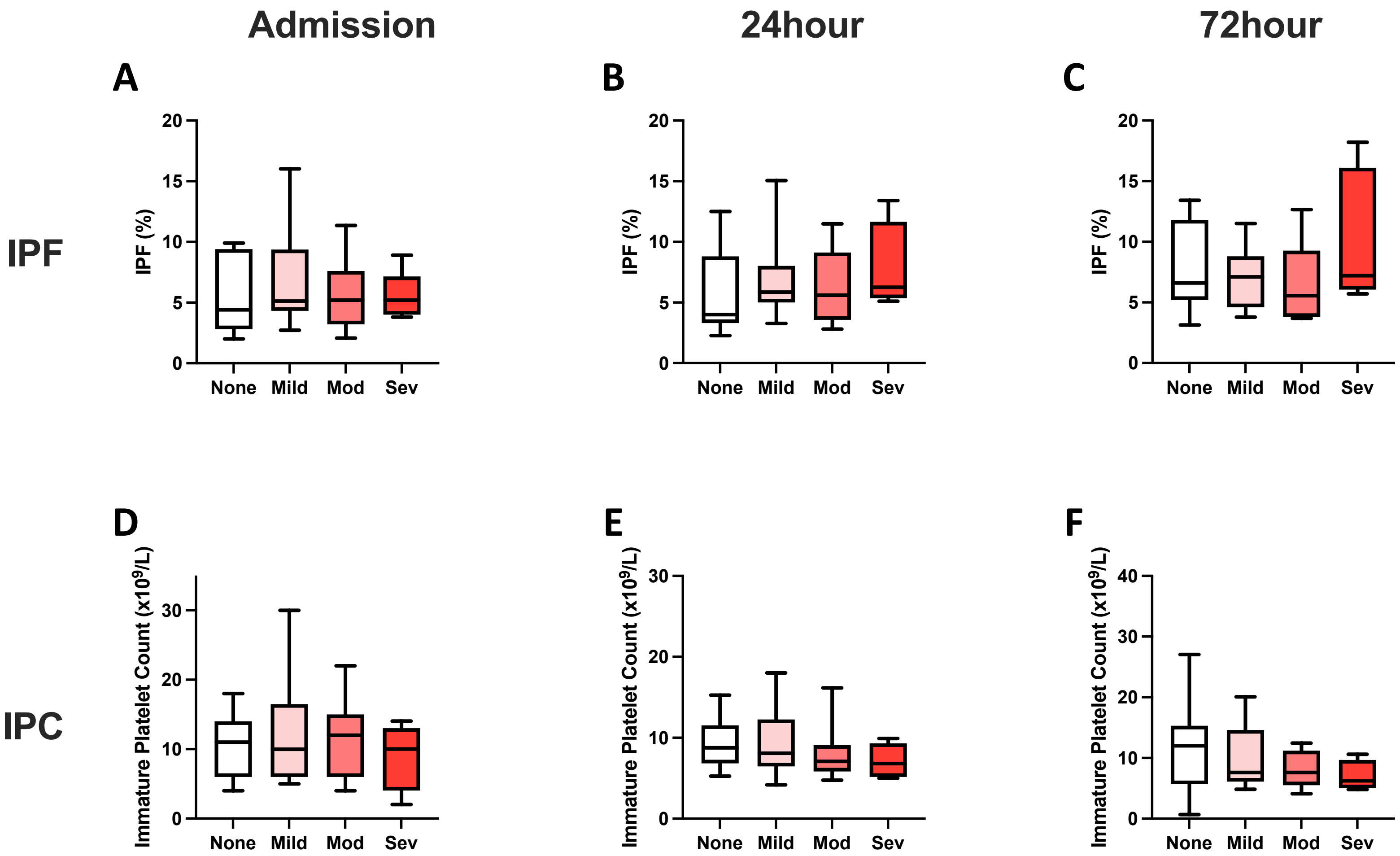

**Supplemental Figure 3.** Organ injury score trajectories stratified by the severity of thrombocytopenia. Values are mean with 95% confidence intervals.

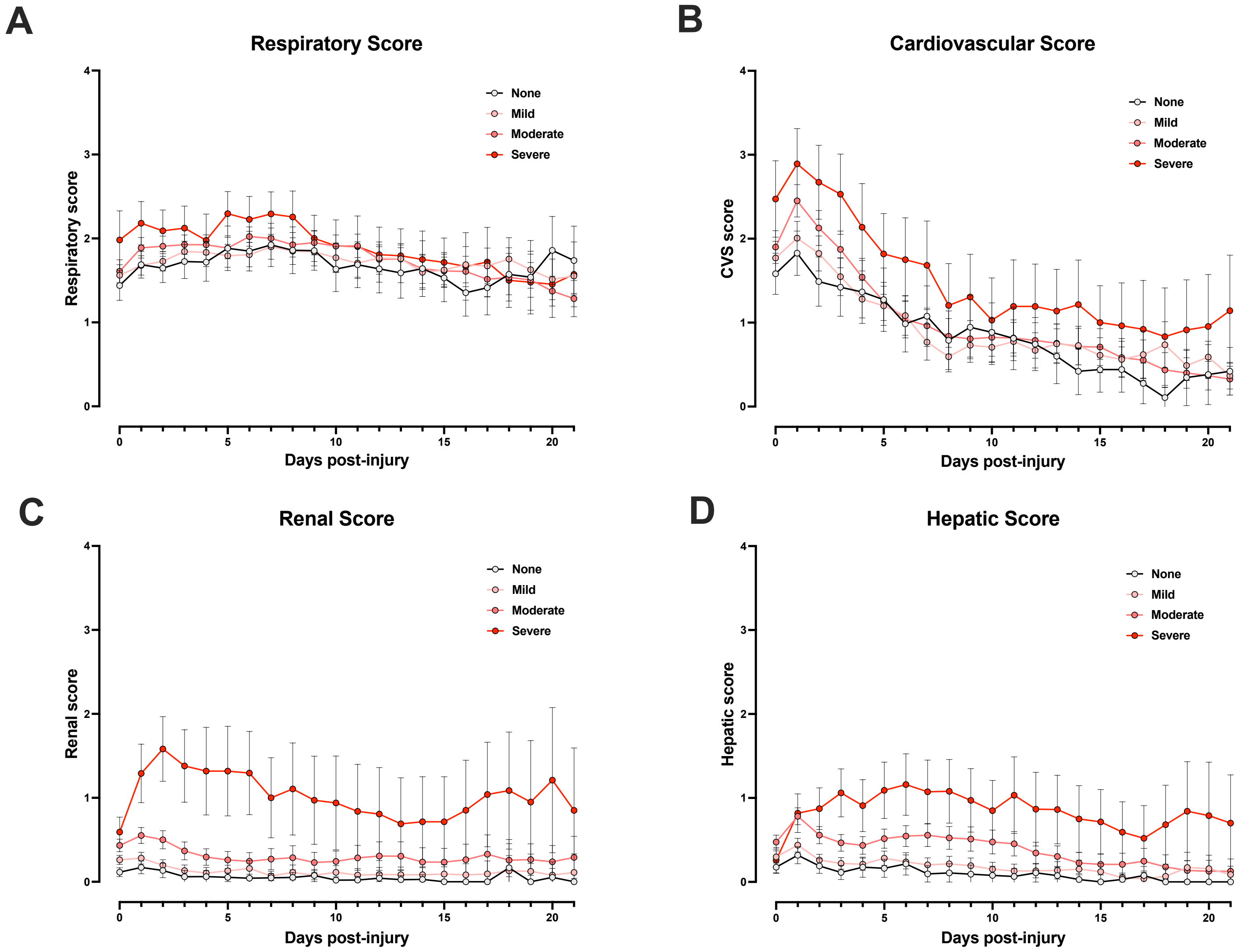

STROBE Statement—Checklist of items that should be included in reports of ***cohort studies***

| | Item No | Recommendation | Page No |
|---|---|---|---|
| **Title and abstract** | 1 | (*a*) Indicate the study's design with a commonly used term in the title or the abstract<br>(*b*) Provide in the abstract an informative and balanced summary of what was done and what was found | 3 |
| **Introduction** | | | |
| Background/rationale | 2 | Explain the scientific background and rationale for the investigation being reported | 4 |
| Objectives | 3 | State specific objectives, including any prespecified hypotheses | 4 |
| **Methods** | | | |
| Study design | 4 | Present key elements of study design early in the paper | 5 |
| Setting | 5 | Describe the setting, locations, and relevant dates, including periods of recruitment, exposure, follow-up, and data collection | 5 |
| Participants | 6 | (*a*) Give the eligibility criteria, and the sources and methods of selection of participants. Describe methods of follow-up<br>(*b*) For matched studies, give matching criteria and number of exposed and unexposed | 5 |
| Variables | 7 | Clearly define all outcomes, exposures, predictors, potential confounders, and effect modifiers. Give diagnostic criteria, if applicable | 5-6-7 |
| Data sources/ measurement | 8* | For each variable of interest, give sources of data and details of methods of assessment (measurement). Describe comparability of assessment methods if there is more than one group | 5-6 |
| Bias | 9 | Describe any efforts to address potential sources of bias | - |
| Study size | 10 | Explain how the study size was arrived at | 5 |
| Quantitative variables | 11 | Explain how quantitative variables were handled in the analyses. If applicable, describe which groupings were chosen and why | 6-7 |
| Statistical methods | 12 | (*a*) Describe all statistical methods, including those used to control for confounding<br>(*b*) Describe any methods used to examine subgroups and interactions<br>(*c*) Explain how missing data were addressed<br>(*d*) If applicable, explain how loss to follow-up was addressed<br>(*e*) Describe any sensitivity analyses | 6-7 |
| **Results** | | | |
| Participants | 13* | (a) Report numbers of individuals at each stage of study—eg numbers potentially eligible, examined for eligibility, confirmed eligible, included in the study, completing follow-up, and analysed<br>(b) Give reasons for non-participation at each stage<br>(c) Consider use of a flow diagram | 8 |
| Descriptive data | 14* | (a) Give characteristics of study participants (eg demographic, clinical, social) and information on exposures and potential confounders<br>(b) Indicate number of participants with missing data for each variable of interest<br>(c) Summarise follow-up time (eg, average and total amount) | 8-9 |
| Outcome data | 15* | Report numbers of outcome events or summary measures over time | 8-9 |

| | | | |
|---|---|---|---|
| Main results | 16 | (*a*) Give unadjusted estimates and, if applicable, confounder-adjusted estimates and their precision (eg, 95% confidence interval). Make clear which confounders were adjusted for and why they were included<br>(*b*) Report category boundaries when continuous variables were categorized<br>(*c*) If relevant, consider translating estimates of relative risk into absolute risk for a meaningful time period | 9-10 |
| Other analyses | 17 | Report other analyses done—eg analyses of subgroups and interactions, and sensitivity analyses | - |
| **Discussion** | | | |
| Key results | 18 | Summarise key results with reference to study objectives | 11 |
| Limitations | 19 | Discuss limitations of the study, taking into account sources of potential bias or imprecision. Discuss both direction and magnitude of any potential bias | 12 |
| Interpretation | 20 | Give a cautious overall interpretation of results considering objectives, limitations, multiplicity of analyses, results from similar studies, and other relevant evidence | 12 |
| Generalisability | 21 | Discuss the generalisability (external validity) of the study results | 12 |
| **Other information** | | | |
| Funding | 22 | Give the source of funding and the role of the funders for the present study and, if applicable, for the original study on which the present article is based | 1 |

*Give information separately for exposed and unexposed groups.

**Note:** An Explanation and Elaboration article discusses each checklist item and gives methodological background and published examples of transparent reporting. The STROBE checklist is best used in conjunction with this article (freely available on the Web sites of PLoS Medicine at http://www.plosmedicine.org/, Annals of Internal Medicine at http://www.annals.org/, and Epidemiology at http://www.epidem.com/). Information on the STROBE Initiative is available at http://www.strobe-statement.org.
